# Artificial intelligence in the respiratory sounds analysis and computer diagnostics of bronchial asthma

**DOI:** 10.1101/2021.11.18.21266503

**Authors:** A. Gelman, V. Sokolovsky, E. Furman, N. Kalinina, G. Furman

## Abstract

Using a database containing audio files of respiratory sound records of asthmatic patients and healthy patients, a method of computer-aided diagnostics based on the machine learning technique – creation of neural networks, has been developed. The database contains 952 records of respiratory sounds of asthma patients at different stages of the disease, aged from several months to 47 years, and 167 records of volunteers. Records were carried out with a quiet breathing at four points: in the oral cavity, above the trachea, on the chest, the second intercostal space on the right side, and at a point on the back.

The developed method of computer-aided diagnostics allows diagnosing bronchial asthma with high reliability: sensitivity of 89.3%, specificity of 86%, accuracy of about 88% and Youden’s index of 0.753.

The program learned once makes it possible to diagnose bronchial asthma with high reliability regardless of patient’s gender and age, a stage of disease, as well as the point of sound recording.

The developed method can be used as an additional screening method for the diagnostics of bronchial asthma and serve as the basis for development of computer control methods, including remote control (telemedicine) of patient’s condition and the effectiveness of the applied drugs in real time.

## Introduction

Diagnostics of bronchial asthma is based on a comprehensive dynamic examination of a patient, including determination of the functional state of the lungs, etc. [1]. One of the main tasks of asthma treatment is early diagnosis and timely initiation of treatment, as well as achievement of control over the patient condition, prevention of exacerbations of illness and occurrence of severe forms of disease that requires a constant control including remote out-of-hospital monitoring [1]. No less important is a quick and objective determination of the effectiveness of the prescribed drugs for a given patient. Determination of the lungs functional state is difficult in some cases of pediatric practice (preschool children insufficiently cooperate during the examination), and the results of physical examination by auscultation of the lungs by a doctor are largely subjective, and sometimes timely auscultation of the lungs by a doctor is delayed, as, for example, it occurs during the COVID-19 pandemic [2,3].

Computer-aided analysis of respiratory sounds can become an additional screening method for diagnosing pulmonary diseases, including bronchial asthma in children [4,5]. Computer-aided methods for diagnosing pulmonary diseases are devoid of subjectivity and allow analyzing changes in respiratory sounds, which cannot be detected by the human ear. Changes in the characteristics of the airways caused by various diseases lead to appearance of additional pathological noises, the analysis of which is used in development of computer-aided methods for the diagnostics of pulmonary diseases [6,7,12]. Respiratory sounds, accompanying broncho-obstructive syndrome can be characterized by the appearance / amplification of a set of periodic waves with a fundamental frequency in the range from 100 Hz to 2500 Hz. In works [8,9] it is indicated that the fundamental frequency lies between 100 Hz and 1000 Hz and between 400 Hz and 1600 Hz, respectively. At the same time, the dominant frequency of wheezing is above 400 Hz, while of wet wheezing it is about 200 Hz and less [9, 10]. The breathing of patients with asthma is characterized by wheezing sounds in the expiratory phase, the wheezing duration is in the range from 80 ms to 250 ms. Using these features of respiratory sounds of patients with asthma, the methods of computer-aided diagnostics of broncho-obstructive syndrome in children were developed [4,5,11]. These methods are based on an automated comparison of the Fourier spectra of respiratory sounds of the healthy and asthmatic children and demonstrate high diagnostic values (AUV varies from 0.783 to 0.895). One cannot exclude variability of this sound phenomenon depending on the personal characteristics of a patient with bronchial asthma (age, sex, stage of disease, individual characteristics of physical development).

The use of modern electronic devices (computers, mobile phones, etc.), communication facilities, and software makes it is possible to develop new methods of computer-aided diagnostics based on the analysis of patients’ breathing sounds. These methods include machine learning methods and deep machine learning methods [12,13]. However, the development of such diagnostic methods requires the analysis of a large database with the maximum possible number of patients and volunteers.

In this work, machine learning methods were used to develop a computer-aided method for diagnostics of bronchial asthma. The method is based on a comparison of respiratory sounds of patients with bronchial asthma and healthy volunteers. The database created by us contains over 1000 respiratory sound records (see the next section).

### Database

The anonymous database, along with digital audio files containing recordings of respiratory sounds, covers the main characteristics of the examined person: age, sex, health information (healthy, exacerbation of bronchial asthma or its remission), recording time, recording point (see below). The database contains 952 recordings of respiratory sounds of asthma patients whose age is from several months to 47 years and 167 records of healthy volunteers in the same age range. The records are distributed among men and women as 67% and 33%, respectively.

Clinical examination of patients and recording of respiratory sounds were performed at the Regional Children’s Clinical Hospital in Perm (Russia). The examinations were carried out in accordance with the Declaration of Helsinki, adopted in June 1964 (Helsinki, Finland), revised in October 2000 (Edinburgh, Scotland), and approved by the Ethics Committee of E.A. Vagner Perm State Medical University. The written informed consent was obtained from the examined persons (at examination of children - from their parents or guardians) in accordance with the Federal Law “Fundamentals of the Legislation of the Russian Federation on the Protection of Citizens’ Health” from July 22, 1993, No. 54871.

Bronchial asthma in patients was diagnosed in accordance with the recommendations presented in the National Program “Bronchial Asthma in Children. Treatment Strategy and Prevention” [14]. According to a comprehensive medical examination, 232 records were made for patients in a stage of exacerbation, 309 for patients in a stage of remission, and 410 in a stage of incomplete remission. Auscultatory data in patients with bronchial asthma were characterized by hard breathing and presence of diffuse dry wheezing over both lungs. Vesicular breathing was heard over the lungs of healthy volunteers with good conduction of sound to all parts of the lungs. At the time of recording, the volunteers were not suffering from any pulmonary diseases or other diseases causing pathological changes in respiratory sounds.

### A. Respiratory sound registration

Respiratory sounds were recorded at four points during quiet breathing: in the oral cavity (point 1, 4.3% of all records), above the trachea (point 2, 50.2%), on the chest, the second intercostal space on the right side, (point 3, 23.6%) and at the point on the back (point 4, 21.9% of all records). For most of the persons examined, the recordings were performed at several points and for some patients also at different stages of the disease: exacerbation, incomplete remission and remission. The recording systems and the quality of the records met the requirements formulated [11]. The quality of the recordings was controlled visually (oscillograms of the recordings were shown on a computer screen) and using the program developed by us.

Respiratory sounds were recorded continuously during several respiratory cycles, approximately for 25 s. It allows us to reduce the effect of random variations of the sound intensity on the analysis results.

Respiratory sounds were recorded using mobile phones and computer systems. The audio files recorded with the help of the phones using both the built-in and the external microphones, were sent to the “cloud” to create an anonymous database. The computer recordings were performed using the developed computer-aided systems for recording respiratory sounds [4,5,7,11], external microphones, electronic phonendoscopes and computer sound cards, Adobe Audition system. All systems demonstrated high amplitude-frequency linearity over a frequency range of 100 Hz to 3000 Hz. The sampling rate varied from 22 kHz to 96 kHz.

### Method of data processing

The developed methods of computer-aided diagnostics of pulmonary diseases are usually based on analysis of respiratory sound oscillograms, their spectrograms or Fourier spectra [2-11,15,16]. An alternative approach could be the following: to develop diagnostic methods based on machine learning and deep machine learning techniques. Recently, the studies of the possibility of applying such methods in various fields of medicine have been actively carried out (see [12,13,17-21] and references therein). At present, there is a large number of machine learning methods that allows creating decision-making algorithms suitable for a given situation and able to solve various tasks, including medical ones (e.g., classification of various types of cancer [18], analysis of the effectiveness of dialysis [18], diagnosis of pulmonary diseases [12,13,20,36]). Machine learning methods are proposed to be widely used, in particular, in pulmonology: for sorting patients with chronic obstructive pulmonary disease [21], for automatic classification of respiratory sound spectra in patients with pulmonary diseases [22], for diagnosis of bronchial asthma and chronic obstructive pulmonary disease [23] and ranking of clinical assessments in children with asthma [24].

Among the tasks of machine learning, three main ones can be highlighted: the problems of classification, clustering, and regression [25-29]. At solution of the regression problems, the mathematical relationship between a function and an argument (or arguments) is restored. The solution of the clustering problem, which refers to automatic learning, without user / programmer interference (unsupervised learning), it is designed to divide the data into certain categories according to the selected common features. When solving clustering problems, the number of categories, which will be obtained as a result, is unknown. In the case of automatic diagnostics, it is necessary to solve classification problems, which are similar in formulation to clustering problems, but at the same time, the number of categories is known in advance. The main problem comes down to choosing the most efficient algorithm, which allows achieving the maximum accuracy for the test dataset. The purpose of solving classification problems is to assign a phenomenon or parameter to a specific category, in our case, to establish the correspondence of the respiratory sound to a healthy volunteer or a patient with bronchial asthma. The difficulty in choosing an algorithm lies in the fact that there is a large number of different algorithms for solving clustering problems, and each of them is better suited for some particular class of problems.

Our analysis of the methods showed that two methods can be used with great efficiency to classify audio recordings: analysis using a neural network [29] and analysis using a method called a solution tree [25]. Several solution trees can be combined into a so-called random forest [20], which has an extra accuracy compared with a single solution tree, since the final result in the case of classification problem is determined by the “voting” of all trees which make up the random forest. In some cases, the use of the random forest method for classification gives a higher accuracy in comparison with a neural network, but the advantage of a neural network over the random forest method is that with each incorrect solution of the classification problem, the neural network can be additionally trained under human supervision.

Mathematically, a neural network consists of a set of functions (neurons), each of which possesses its own coefficient of connection with other functions [26]. Thus, when an input signal comes to a neuron, as a result of simple mathematical operations, a decision is made which neuron will be the next to be activated. Thus, the procedure reaches the last stage (the last neuron), the result of which will coincide with one of the categories into which the input data should be divided as a result of solving the problem. In our case, the input data are the digital recordings of respiratory sounds. The recordings are divided into two categories at the output: healthy and sick. In addition to the above noted advantages of the neural networks over the random forest method, it is worth mentioning that a trained neural network can fairly quickly work on relatively low-power computers [27]. The latter advantage of networks is important for development of methods of computer-aided diagnostics in real time, remote screening diagnostics and monitoring of the patient condition, as well as for development of outside medical diagnostics and condition monitoring (for example, parental control over the child condition; the result of such monitoring should be recommendation to visit a doctor in time).

Taking into account the above, we have chosen the neural network method. To process the respiratory sound audio files, the program based on the Python language, version 3.8, using the Keras and Librosa libraries was developed. The first library is used to build a neural network, the second is used for simple and quick analysis of audio files. The time required to analyze an audio file is of the order of a second. To build the neural network, a sequential model was chosen from the standard set of the Keras library. It allows creating a network of several layers, train it and get a forecast for the category (define the category), to which the object should belong (audio file), the characteristics of which are supplied to the input. A neural network consists of at least two layers of the so-called neurons, which are the functions, processing the parameters of the analyzed object and activating the element of the next layer depending on the result of work of the previous layer [25-29].

The developed method is based on the following: the audio recording of respiratory sounds is converted into a time series, the characteristics of which can be investigated by any methods suitable for time series analysis. These characteristics can subsequently be used to train the neural network and develop methods for express analysis. The Fourier spectrum of respiratory sounds of asthmatic patients contains frequency ranges, in which the amplitudes of harmonics significantly exceed the amplitudes in the spectrum of healthy patients (see, for example, Fig. 4 in [3] and Fig. 3 in [10]). There is a periodic manifestation of such an increase in time. The duration of such amplifications is about 200 ms (see, for example, Fig. 5 in [3]). This allows one to visually distinguish the respiratory sounds of sick and healthy persons. Note that asthmatic disease is often accompanied by an increase in respiratory rate. These differences between the characteristics of the respiratory sounds of the patients and the healthy volunteers allowed us to choose the following parameters for training the neural network: spectral bandwidth, spectral centroid, zero-crossing rate, spectral roll-off and chromaticity. Our results have shown that a set of these parameters is sufficient for constructing and training a network, application of which makes it possible to achieve a high level of reliability of computer-aided diagnostics.

A spectral width is determined as a width of the spectrum at half its maximum intensity and shows how much the energy is concentrated or dispersed depending on the frequency.

The spectral centroid determines position of the energy center of the spectrum and is defined as

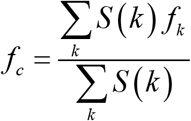

where *S* (*k*) is the amplitude of the *k* -th harmonic with the frequency *fk*.

The number of zero crossings characterizes the sound recording in terms of the presence or absence of high frequency harmonics. Mathematically, it shows how smooth the signal is.

A spectral roll-off is a measure of the waveform that indicates the frequency after which the amplitudes of the high frequency harmonics drop to zero.

A chroma is a vector usually containing 12 elements. To determine the vector elements, the spectrum is divided into 12 sections (classes) and an element is determined by the amount of energy in the corresponding class. Chroma is used to describe the measure of similarity between signals.

To calculate the parameters noted above, the built-in functions of the Librosa library (chroma_stft, spectral_centroid, spectral_bandwidth, spectral_rolloff, zero_crossing_rate) were used. The functions allow one to calculate these parameters quickly; the records contain up to a million points and use of other software can lead to a significant increase in the calculation time. In the standard library, these functions are optimized and run as fast as possible.

The database of respiratory sound recordings was divided into two groups: the first, training group, which contains 374 records of respiratory sounds of asthmatic patients and 146 records of healthy volunteers, was used to train the network; the second, control one (577 - sick and 21 - healthy), was used to check the operation of the network and the correctness of computer-aided diagnostics. In these groups, the examined persons (their records) were almost equally distributed by age, gender, and recording points. None of the recordings included in the first group was used in the second one. The performed separation is dictated, on the one hand, by the need to “balance” the training group (it is desirable that the numbers of recordings of patients and of healthy volunteers were equal), on the other hand, the number of recordings used to train the network should be large (at least several hundred). In this work, when training the network and checking its operation, there was no division of patients according to the disease stages. To demonstrate the universality of the developed approach, the training group contained recordings made on different devices and at different recording rates from 44 kHz to 96 kHz, the control group - only records performed using a computer at a rate of 22 kHz.

Computer analysis of respiratory sounds was carried out at the Ben-Gurion University (Israel). The developed software based on the Python scripting language was used. TKinter library was used as a graphical interface (GUI) convenient for a user.

## Results

The developed program correctly diagnosed “a patient with bronchial asthma” in 516 cases out of 577 ones, 55 recordings were mistakenly identified as recordings of healthy persons, and in 6 cases diagnosis was not unambiguously determined. The diagnosis was established correctly for 18 volunteers from 21.

The results allow us to assess the main characteristics of the proposed method. The sensitivity of the method is determined by the formula [31]:

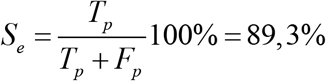

and its specificity as

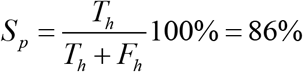

where *T*_*p*_ and *T*_*h*_ are the number of the correctly diagnosed persons as sick and healthy, respectively; *F*_*p*_ and *F*_*h*_ are the number of misdiagnosed patients as healthy and of healthy volunteers as sick, respectively.

The method accuracy is usually determined using the following formula

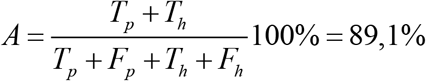

and in our case practically coincides with the sensitivity. This is due to the strong “imbalance” of the recordings number in patients and healthy volunteers in the control group; there are significantly more recordings made for patients, about 30 times, than for healthy volunteers. Approximation for the case of a “balanced” control group (the number of recordings in patients and healthy persons is the same) allows us to estimate the accuracy as 87.5%. To increase the number of recordings in healthy persons in the control group, 50 recordings of healthy persons were randomly selected from the training group. These recordings were added to the control group. This procedure was repeated several times. The spread in specificity for the extended control groups was ±0,1%.

Youden’s index was calculated using the following formula:

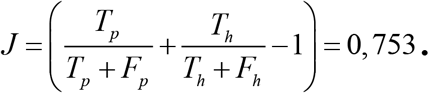

## Conclusion

The proposed method of computer-aided diagnostics, developed using machine learning methods, allows diagnosing bronchial asthma with high reliability: sensitivity of 89.3%, specificity of 86%, accuracy about of 88%, and Youden’s index 0.753. The achieved characteristics of the method exceed those of traditional diagnostic methods of spirometry [32]: sensitivity 29% (when performed by high-level specialists and active cooperation of the patient it can reach 39.8%), specificity 90%; change in the peak flow in the SAPALDIA study [33]: sensitivity 40% and specificity 83%; in the diagnosis of the disease in children [34]: sensitivity 31% and specificity 90%.

The determined parameters of the trained neural network do not depend on the age and gender of the subject, as well as on the recording point. The same parameters can be used in the diagnostics of bronchial asthma disease in different stages (exacerbation, remission, incomplete remission). This can be explained as follows: bronchial asthma is characterized by obstruction and inflammation, which affect all airways from the central to the peripheral parts of the tracheobronchial tree (small bronchi) [35]. These pathological changes in the airways lead to variations in respiratory sounds and changes in the amplitude-frequency characteristic of sounds and to appearance of additional harmonics in the Fourier spectra. The use of drugs reduces inflammation and expands the airways; the patient comes in the incomplete remission or remission stage of the disease. Although at these stages the external signs of the disease (shortness of breath, dyspnea, rapid breathing) may not be observed, lung function is not fully restored that is reflected in the characteristics of respiratory sound. Moreover, the manifestation of pathological processes in respiratory sounds of patients in remission stage may be more pronounced than in the sounds of the patients during exacerbation. Thus, the energy of the pathological respiratory sounds in the remission stage may be higher than the energy during the exacerbation (Fig. 1).

**Fig. 1.**
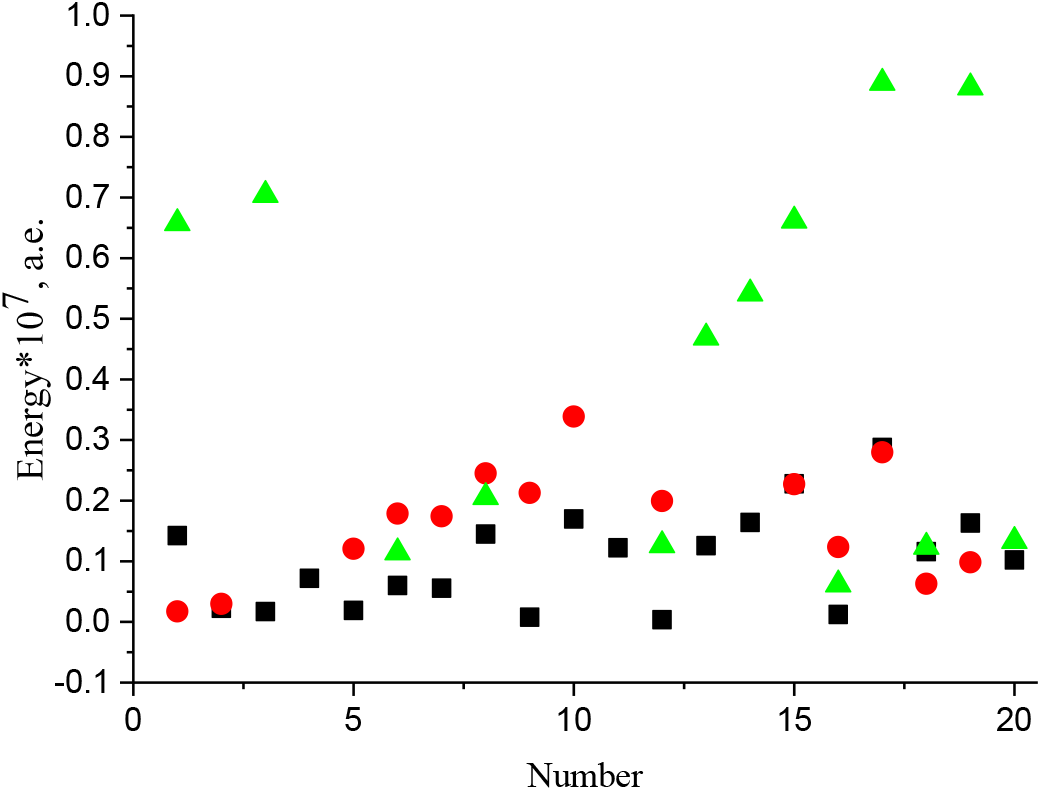
Respiratory sound energy: black squares - for healthy volunteers; red circles - for asthmatic patients in exacerbation stage; green triangles - for asthmatic patients in remission stage. The vertical axis indicates the sound energy determined using the method proposed in [5]. On the horizontal axis a number of the subject is indicated; the numbering of volunteers and patients in both stages is independent. As an example, the results for 20 subjects in each group are presented.

Unlike auscultation of the lungs performed by doctors, the results of computer diagnostics do not depend on the subjective assessment of the doctor, and possess a higher sensitivity when the level of pathological sounds is low and indistinguishable by the human ear against the background of other sounds.

The developed method can be used as an additional express method for screening-diagnostics of bronchial asthma and serve as the basis to develop methods for computer-aided monitoring of the patient condition and the effectiveness of the use of drugs in real time. The proposed method can be used to diagnose and monitor the health state of children under 5 years of age, for whom it is difficult to conduct physical examination and spirometry, to diagnose patients in remote areas and to quickly diagnose patients outside the hospital as well as be applied in telemedicine.

## Data Availability

All data produced in the present study are available upon reasonable request to the authors

## Acknowledgments

This research was supported by a grant from the Ministry of Science & Technology (MOST, N^0^ 3-16500), Israel & Russian Foundation (RFBR), Russian Federation (the joint research project N^0^ 19-515-06001).

## Notes

### Competing Interest Statement

The authors have declared no competing interest.

### Funding Statement

This research was supported by a grant from the Ministry of Science & Technology (MOST, N0 3-16500), Israel & Russian Foundation (RFBR), Russian Federation (the joint research project N0 19-515-06001).

### Author Declarations

The examinations were carried out in accordance with the Declaration of Helsinki, adopted in June 1964 (Helsinki, Finland), revised in October 2000 (Edinburgh, Scotland), and approved by the Ethics Committee of E.A. Vagner Perm State Medical University.

